# Study of SARS-CoV-2 genomes from Pakistan in the post-pandemic period 2023-2024 shows unique phylogenetic diversity as compared with global trends

**DOI:** 10.1101/2025.08.29.25334710

**Authors:** Akbar Kanji, Ali Raza Bukhari, Javaria Ashraf, Amina Ajmal, Maliha Yameen, Sadaf Balouch, Furqan Kabir, Hassan Ghayaz, Muhammad Imran Nisar, Waqasuddin Khan, Zahra Hasan

**Affiliations:** Department of Pathology and Laboratory Medicine, Aga Khan University, Karachi, Pakistan; CITRIC Center for Bioinformatics and Computational Biology, Aga Khan University, Karachi, Pakistan; Department of Pediatrics and Child Health, Aga Khan University, Karachi, Pakistan

**Author notes:** Corresponding author: Zahra Hasan, Department of Pathology and Laboratory Medicine, Aga Khan University, Stadium Road P.O. Box 3500, Karachi 74800, Pakistan.

**Keywords:** SARS-CoV-2, Whole genome sequencing, transmission dynamics, phylogenetics

## Abstract

**Introduction:** SARS-CoV-2 infections caused a global pandemic between 2019 and 2023. Omicron variants continued to emerge but morbidity and mortality were reduced likely due to vaccinations and increasing host immunity. COVID-19 case numbers reported from Pakistan as was genomic epidemiology of SARS-CoV-2. We studied SARS-CoV-2 variants reported between 2023 and 2024 and compare these with variants in other countries.

**Methodology:** We studied SARS-CoV-2 genomes submitted from Pakistan available in public databases as well as those reported by our facility, the Aga Khan University Hospital, Karachi. Phylogenetic analysis was performed. Viral variants reported from other countries were compared.

**Results:** In Pakistan, SARS-CoV-2 genomic epidemiology identified predominant types to be: XBB.1 and its subvariants in 2023 and JN.1 and its subvariants in 2024. Although variants such as KP.2, LB.1, and LA.1 emerged by mid-2024, JN.1 remained the dominant variant in Pakistan. Global data (USA, UK, France, Germany, Italy) showed XBB.1.5 transitioned to JN.1 and KP.3.3 by September 2024. Notably, JN.1 strains persisted in India until 2024.

**Conclusions:** We show differing trends between the evolution of SARS-CoV-2 variants in different countries. Strains in Pakistan differed from four countries in the North. These findings suggest the importance of continued genomic surveillance and epidemiological monitoring to track variant evolution. Different strains likely impact disease severity across regions and may contribute to differences observed in COVID-19 associated morbidity observed globally.

## Introduction

Coronavirus disease 2019 (COVID-19) is caused by the severe acute respiratory syndrome coronavirus 2 (SARS-CoV-2), which was first identified in December 2019 (1). It rapidly evolved into a global pandemic and became the fifth documented pandemic declared by World Health Organization (WHO)(2). Globally, more than 780 million cases and over 7 million deaths have been reported(3). Since the emergence of SARS-CoV-2 in late 2019, the virus has undergone continuous evolution, giving rise to a diverse range of variants with varying transmissibility, immune evasion potential, and disease severity. SARS-CoV-2 circulates widely without clear seasonality, causing both acute infections and long-term post-COVID-19 conditions. It was reported differently across countries depending on the severity of disease as well as the capacity of countries to test, monitor and report SARS-CoV-2 spread. With up to 1 million SARS-CoV-2 diagnostic tests conducted daily in USA during 2021, whilst Pakistan’s testing peaking at approximately 46,500 tests per day in July 2021, when a network of over 110 COVID-19 PCR laboratories was operational(4). SARS-CoV-2 variants are classified by the World Health Organization (WHO) and other national bodies into Variants of Concern (VOCs), Variants of Interest (VOIs), or Variants Under Monitoring (VUMs), based on their potential impact on public health (5). Whole genome sequencing (WGS) was essential for tracking the molecular evolution of SARS-CoV-2, enabling real-time surveillance of emerging variants. However, whilst COVID-19 surveillance continued over the past five years, its effectiveness varied across different regions (6).

In Pakistan reported an estimated 1.57 million COVID-19 cases from 30.6 million SARS-CoV-2 PCR tests (https://ourworldindata.org/coronavirus-testing) with approximately 30,000 deaths have been documented since the onset of the pandemic (7). Several variants, including Alpha (B.1.1.7), Delta (B.1.617.2), and multiple Omicron sub-lineages have been reported in the region [5,6]. However, due to the scarcity of WGS data from Pakistan, SARS-CoV-2 genomic epidemiology from the country has been limited. Pakistan experienced five successive COVID-19 waves were between 2020 and 2022 occurring between; March and July 2020, October 2020 and January 2021, April and May 2021, July and September 2021 and, between December 2021 and February 2022 (8).

In the early pandemic period of 2020 there was limited reporting of strains as they emerged from L, S, V to GH clades (9). In 2021, the emergence of VOC alpha, delta and omicron variants were reported (10). By March 2023, the pandemic was declared to have ended officially. Omicron strains have continued to evolve and according to the WHO COVID-19 epidemiological update, with VOI JN.1, and various VUMs to date. The continuous evolution of SARS-CoV-2 variants has raised concerns regarding vaccine effectiveness associated with Omicron sub-lineages which exhibit varying degrees of immune escape. Vaccination helped manage the COVID-19 pandemic, with introduction and roll out of vaccines in Pakistan from February 2021(11). There is limited information related to the post-pandemic period and the strains circulating 2023 onwards.

This study utilizes whole genome sequencing (WGS) to investigate the genetic evolution of SARS-CoV-2 Omicron variants from January 2023 to September 2024, providing insights into emerging mutations, lineage dynamics, and their potential implications for transmission. We analyzed SARS-CoV-2 genomic data submitted from Pakistan and also that conducted at our institutions. SARS-CoV-2 Omicron lineages were compared with global trends and we also analyzed the temporal distribution and age-associated prevalence of predominant variants. Our data show that there are regional differences in the evolution of SARS-CoV-2 strains globally, this likely has affected the variable morbidity and mortality caused by them globally.

## Material and Methods

This study was approved by the Ethical Review Committee (ERC), The Aga Khan University (AKU), Pakistan (ID 2022-6871-22433).

## Data availability

SARS-CoV-2 genome data from Global Initiative on Sharing All Influenza Data (GISAID) was selected for download for the study period January 2023 and September 2024. Datasets were downloaded on 01/01/2025. Data for Pakistan we downloaded data for those with “high coverage” as part of the filtering criteria (https://gisaid.org/). In total, 935 sequences were downloaded.

For global comparative analysis, SARS-CoV-2 genomic surveillance data were retrieved from the GISAID EpiCoV database for six countries: United States, United Kingdom, France, Germany, Italy, and India. For each country, the top 20 most frequently reported lineages during the study period were selected. The total number of sequences retrieved per country were as follows: United States (n = 286,135), United Kingdom (n = 54,574), France (n = 32,137), Germany (n = 22,451), Italy (n = 12,509), and India (n = 14,734).

### SARS-CoV-2 testing at AKUH

We conducted a retrospective review of 282 nasopharyngeal swab samples tested positive for SARS-CoV-2 through reverse transcription (RT) polymerase chain reaction (PCR) using the Cobas SARS-CoV-2 test, Roche Cobas 6800 system at AKU Hospital (AKUH) Clinical Laboratories are accredited by the College of American Pathologists (CAP), USA. Samples included in the analysis were from all age groups and had a CT (crossing threshold) value ≤ 35.

### SARS-CoV-2 WGS at AKU

SARS-CoV-2 PCR positive samples (n=223) which met the sequencing criteria of a CT value ≤26 were processed for WGS as described previously (10). Briefly, RNA was transcribed to complementary DNA (cDNA) was synthesized using the Superscript IV VILO™ master mix (Thermo Scientific, MA, USA) following the guidelines provided by the manufacturer(12). Briefly, cDNA was used as a template and two separate PCR were set up and mixed with Q5 High Fidelity master mix along with specifically designed Primer Pool A and Pool B by ARTIC nCoV 2019 network_V3 by IDT (USA)(13). Samples were prepared and run according to the Illumina Library Prep protocol(14) and sequenced on a MiSeq Platform (Illumina, Portland, USA).

### Phylogenetic analysis of SARS-CoV-2

We conducted phylogenetic analysis on genomes (n=935) from Pakistan which were “complete”, whilst “low coverage” were excluded (**S Table 1**). FASTA files of Omicron sequences from AKUH that met quality metrics (WGS coverage, QC score >70) from used for phylogenetic analysis together with corresponding metadata (age, gender, date of collection and location) using the augur pipeline (15). Full length SARS-CoV-2 genomes of Omicron subvariants were aligned using the MAFFT alignment tool (16). Multiple Sequence Alignment (MSA) files generated from MAFTT were used for a maximum likelihood (ML) phylogenetic tree through IQ-TREE2 (17). By applying a generalized midpoint rooting strategy, branch length variance was calculated using Tree-Time (18). The tree was visualized and edited in Figtree *v*. 1.4.4 (https://tree.bio.ed.ac.uk).

The final tree with annotated nodes and metadata was exported to the phylodynamic visualizing tool Auspice (19).

## Statistical analysis

Demographic results are presented in mean ± SD. Kruskal–Wallis tests was used to analyze statistical significance, with p-value less than 0.05 considered statistically significant. Ms Excel and Graph Pad Prism *v.* 5.0 (http://www.graphpad.com) was used for statistical analysis.

## Results

### SARS-CoV-2 genomic epidemiology in Pakistan

The period of study from January 2023 to September 2024 was near to the start of the official ‘end’ of the pandemic declared in March 2023(20). However, COVID-19 infections persisted albeit testing and reporting was more limited. According to the Our World in Data, in January 2023, the new SARS-CoV-2 cases were (n=584), followed by February 2023 (n=726) and March 2023 (n=2558). From January to February 2023, Pakistan experienced very low new infections (<800/month), followed by a rise to 2,558 new cases in March 2023. This showed a minor resurgence, probably associated with emerging variants. However, the detailed monthly data beyond March 2023 are limited in publicly available sources like Worldometer (21).

To understand the SARS-CoV-2 strains circulating in we used available data for 935 genomes present in GISAID for the period studied. Analyzing this identified a shift in the prevalence of major SARS-CoV-2 variants from XBB through to JN.1 and then LB and KP variants as depicted in (**Fig 1**). The highest number of genomes available were from March 2023 (n=343), followed by March 2024 (n=109). A total of 86 unique variants were identified. Among these, 13 variants showed a relative frequency of ≥20%, including FL.1, FY.5, GW.5, GW.5.1, HK.3, JN.1, JN.1.16, JN.1.18, KP.2.3, KS.1, LA.1, XBB.1.19.1, and XBB.1.9. The remaining 73 SARS-CoV-2 variants occurred at frequencies below 20% (**S1 table**).

**Fig 1:**
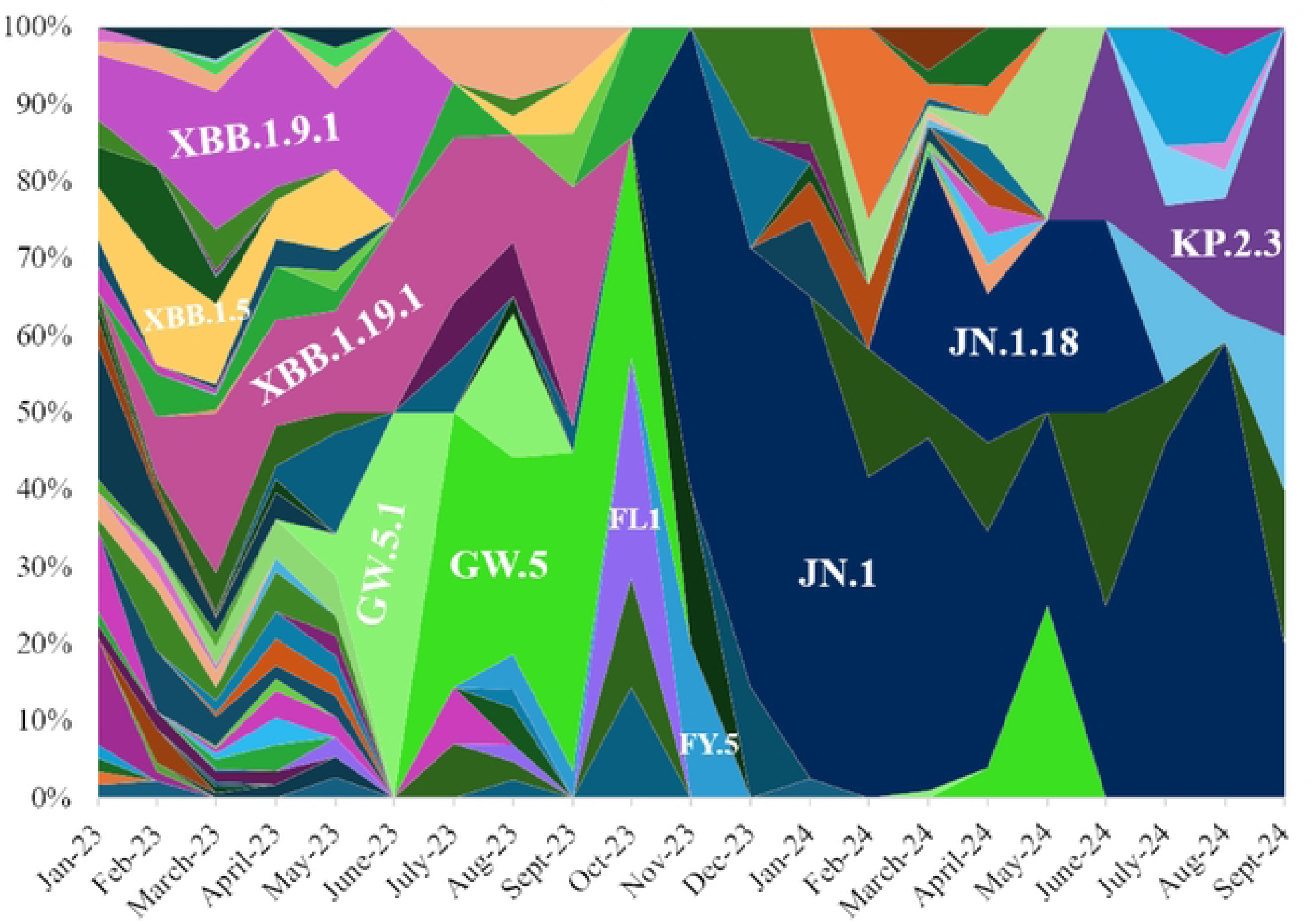
Depicts temporal distribution of 10 major variants of the 86 SARS-CoV-2 variants and sub variants based on GISAID Pakistan data (n=935 samples) from January 2023 to September 2024. Each major variant is represented by a distinct color, illustrating the changing prevalence of variants over time.“

Monthly trends revealed that in March 2023 (n = 343), the dominant variants were XBB.1.19.1 (21%) and XBB.1.9.1 (18%). In April 2023, XBB.1.19.1 accounted for 14% and XBB.1.9.1 for 21%. By August 2023 (n = 43), GW.5 (26%), GW.5.1 (19%), and XBB.1.19.1 (14%) were predominant. In March 2024 (n = 109), the majority of cases were due to JN.1 (46%) and JN.1.18 (31%). Although variants such as KP.2, LB.1, and LA.1 emerged by mid-2024, JN.1 remained the dominant circulating variant by September 2024 (**S1 table**)

### Description of SARS-CoV-2 genomes sequenced at AKU

We wanted to understand the demographic trends associated with COVID-19 Omicron infections to understand SARS-CoV-2 infections caused by different lineages and sublineages during the period. As such data is not available in GISAID, we focused on the genomes sequenced at our own center. The AKU Hospital Clinical Laboratory is based in Karachi but also receives samples for COVID-19 testing from all over the country. A total of 282 RT-PCR SARS-CoV-2 positive samples were reported between January 2023 to September 2024.

Age-wise analysis revealed that adults aged 19–50 years accounted for 54% of cases, followed by older adults >50 years (44%) and children 1–18 years (3%) (**Table 1**). No significant age-wise differences were observed between SARS-CoV-2 specimens studied or between cases in from females or males. The majority of samples (95%) were collected from outpatients, with no significant differences in variant prevalence between inpatient and outpatient settings (**Table 1**).

**Table 1.** Demographcis of SARS-CoV-2 Main Variants and Sub-variants Detected During the period January 2023 to September 2024.

| Demographics | Total Number of Samples (n=192) | XBB.1 (n=55) | XBB.1.5 (n=15) | XBB.1.9 (n=26) | FL (n=25) | GW (n=16) | EG (n=11) | XBB.1.16 (n=3) | JN (n=10) | HK (n=2) | LA.1 (n=6) | LB (n=8) | Recombinant (n=15) | P value |
| --- | --- | --- | --- | --- | --- | --- | --- | --- | --- | --- | --- | --- | --- | --- |
| Gender-wise distribution |  |  |  |  |  |  |  |  |  |  |  |  |  |  |
| Male | 88 (46) | 26 (30) | 6 (7) | 13 (15) | 10 (11) | 8 (9) | 5 (6) | 2 (2) | 5 (6) | 1 (1) | 3 (3) | 2 (2) | 7 (8) | p=0.9537 |
| Female | 104 (54) | 29 (28) | 9 (9) | 13 (13) | 15 (14) | 8 (8) | 6 (6) | 1 (1) | 5 (5) | 1 (1) | 3 (3) | 6 (6) | 8 (8) |  |
| Median Age | 42 (22) | 38 | 36 | 37 | 38 | 42 | 37 | 60 | 50 | 66 | 59 | 46 | 43 |  |
| Age Interquartile Range (25th and 75th percentile) | 28-64 | 27-63 | 27-62 | 28-61 | 27-62 | 28-64 | 27-61 | 29.5-63.5 | 35-73 | 62-66 | 37-72 | 26.75-67 | 29-64 |  |
| Age-wise distribution |  |  |  |  |  |  |  |  |  |  |  |  |  |  |
| 1-18 | 5 (3) | 1 (20) | 0 | 2 (40) | 1 (20) | 1 (20) | 0 | 0 | 0 | 0 | 0 | 0 | 0 | p=0.2943 |
| 19-50 | 103 (54) | 29 (28) | 9 (9) | 15 (15) | 14 (14) | 6 (6) | 5 (5) | 3 (3) | 6 (6) | 0 | 2 (2) | 3 (3) | 11 (11) |  |
| >50 | 84 (44) | 25 (30) | 6 (7) | 9 (11) | 10 (12) | 9 (11) | 6 (7) | 0 | 4 (5) | 2 (2) | 4 (5) | 5 (6) | 4 (5) |  |
| Sample Collection |  |  |  |  |  |  |  |  |  |  |  |  |  |  |
| Karachi |  |  |  |  |  |  |  |  |  |  |  |  |  |  |
| Inpatients | 9 (5) | 1 (11) | 0 | 1 (11) | 1 (11) | 1 (11) | 1 (11) | 0 | 0 | 0 | 2 (22) | 0 | 2 (22) | p=0.145 |
| Outpatients | 183 (95) | 54 (30) | 15 (8) | 25 (14) | 24 (13) | 15 (8) | 10 (5) | 3 (2) | 10 (5) | 2 (1) | 4 (2) | 8 (4) | 13 (7) |  |

Among children aged 1–18 years, infections were recorded in March 2023 (3 cases) and July 2024 (2 cases). In the 19–50 years age group, the highest number of cases was observed in March 2023 (68 cases), followed by a smaller increase in July 2024 (8 cases). We determined the number of COVID-19 cases in each age group for whom viral genome data was available. In individuals aged above 50 years, cases fluctuated over time, with notable peaks in March 2023 (39 cases) and February 2024 (12 cases). Additional cases were reported in January 2023 (9 cases), May 2023 (7 cases), July 2023 (7 cases), May 2024 (4 cases), and July 2024 (10 cases). Overall, most cases were observed in adults and older individuals, with peaks in March 2023 and February 2024 **(Fig 2).**

**Fig 2:**
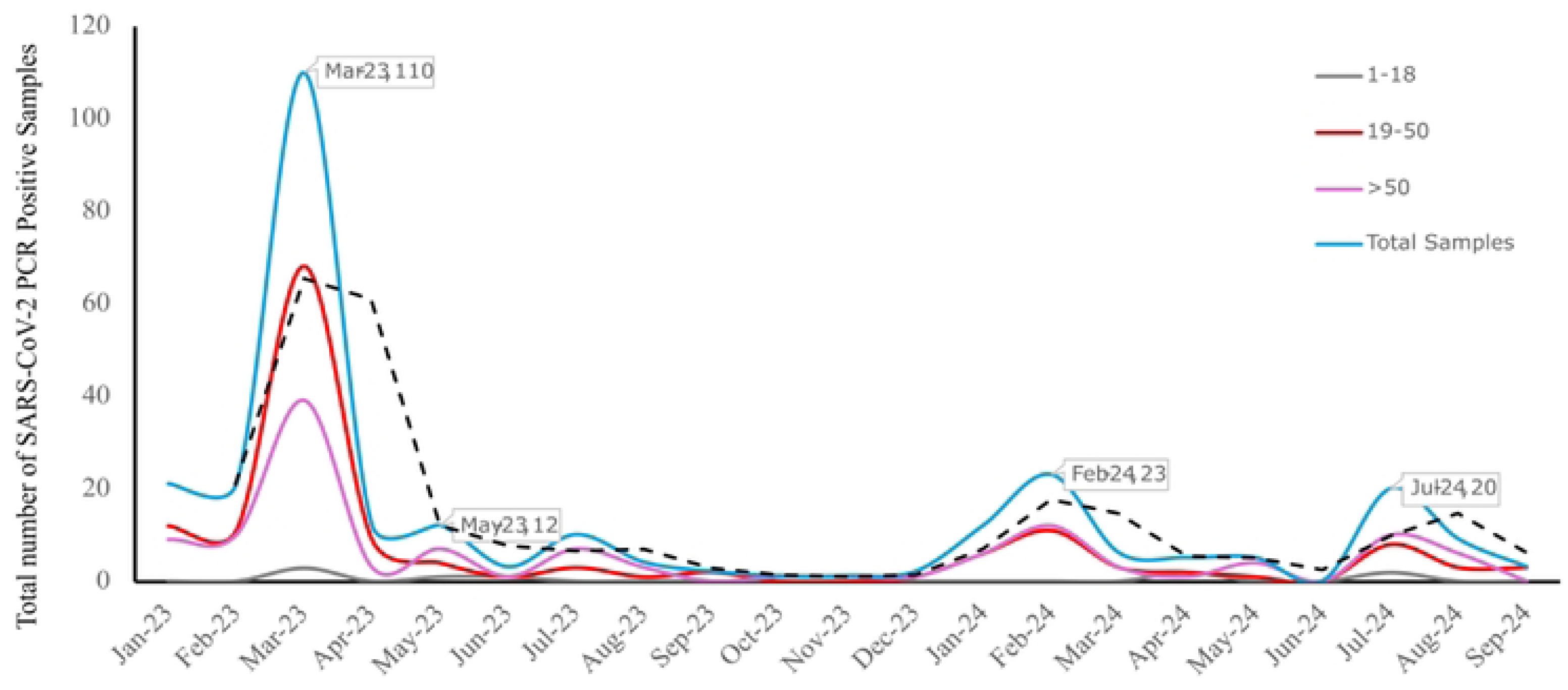
Monthly trend of SARS-CoV-2 positive samples (N=282) across three age groups. The age groups are represented by distinct colors: 1–18 years (grey), 19–50 years (red), and >50 years (pink), The total number of samples is represented by blue color.

### Genomic epidemiology of SARS-CoV-2 at AKU

We performed the whole genome sequence analysis of the 223 Omicron genomes available with their date and location of sample collection from Aga Khan University Hospital (AKUH). We studied the SARS-CoV-2 genome distributions from different provinces of Pakistan. Most were from Karachi, Sindh (94.17%) with limited numbers from Islamabad Capital Territory (ICT), 1.79%; Gilgit Baltistan (GB), 1.35%; Baluchistan and Punjab, 0.9%; Azad Jammu and Kashmir (AJK) and Khyber Pakhtunkhwa (KPK), 0.45% (**S1 Fig**).

WGS revealed that 192 SARS-CoV-2 genomes comprised of twelve major variants; XBB.1 (28.6%), XBB.1.9 (13.5%), FL (13%), XBB.1.5 (7.8%), GW (8.3%), EG (5.7%), and recombinant strains (7.8%) of the total genomes (**Table 1**). Gender-wise distribution showed that XBB.1 was the most prevalent variant among both males (30%) and females (28%), followed by XBB.1.9 (15% in males and 13% in females), and FL (14% in females). There was no statistically significant association between variant distribution and gender (p= 0.9537). XBB.1 was dominant among both adult groups, whereas XBB.1.9 was predominant in children (40%).

### Shift in Omicron variants 2023-2024

We analysed the range of SARS-CoV-2 genomes from the study period and found these to comprise 66 variants and subvariants, distributed across 14 distinct clades: 22F, 22D, 22E, 23A, 23B, 23C, 23D, 23E, 23F, 23H, 24A, 24E, 24G, 24H, and two recombinant groups (XCC and XDK.1.2). Clade 22F included XBB.1 and its sub lineages (XBB.1.48, XBB.1.19.1, XBB.1.11.1, XBB.1.22.1), along with the GW lineage group (GW.3 through GW.5.2) and XBB.2.4. Clade 22D comprised BN.1.3 and BN.1.4, and clade 22E included BQ.1 and BQ.1.1. Clade 23A featured XBB.1.5 sub-lineages (XBB.1.5.9, XBB.1.5.15, XBB.1.5.24) and GV.1. Clade 23B included XBB.1.16.16 and XBB.1.16.8; clade 23C contained CH.1.1.1. Clade 23D included FL (e.g., FL.3.2, FL.4, FL.5, FL.10, FL.12), EG variants (e.g., EG.4, EG.14), and XBB.1.9 sub-lineages (XBB.1.9.1, XBB.1.9.2). Clade 23E included XBB.2.3, and clade 23F included HV.1 and HK.28–29. Clade 24A contained the JN lineage family (JN.1, JN.1.1, JN.1.39, JN.1.61, JN.1.64), LA.1, LB.1, LE.1.2, MA.1, and KS.1. Clades 24E, 24G, and 24H contained KP.3.1.1, KP.2.3 (and KP.2.3.12), and LF.7.1, respectively. Overall, the main variants were: XBB.1 (25%), XBB.1.9 (12%), FL (11%), GW (7%), XCC (7%), XBB.1.5 (7%), EG (5%) and JN.1 (5%). Other lineages identified were LA.1 (3%), LB.1 (4%), KP.2.3 (1%), XBB.2.3 (2%), XBB.1.16 (1.4%), Outgroup (B) (2%). The remaining (<1%) lineages comprise 8% of total genomes analyzed (**Fig 3**).

**Fig 3:**
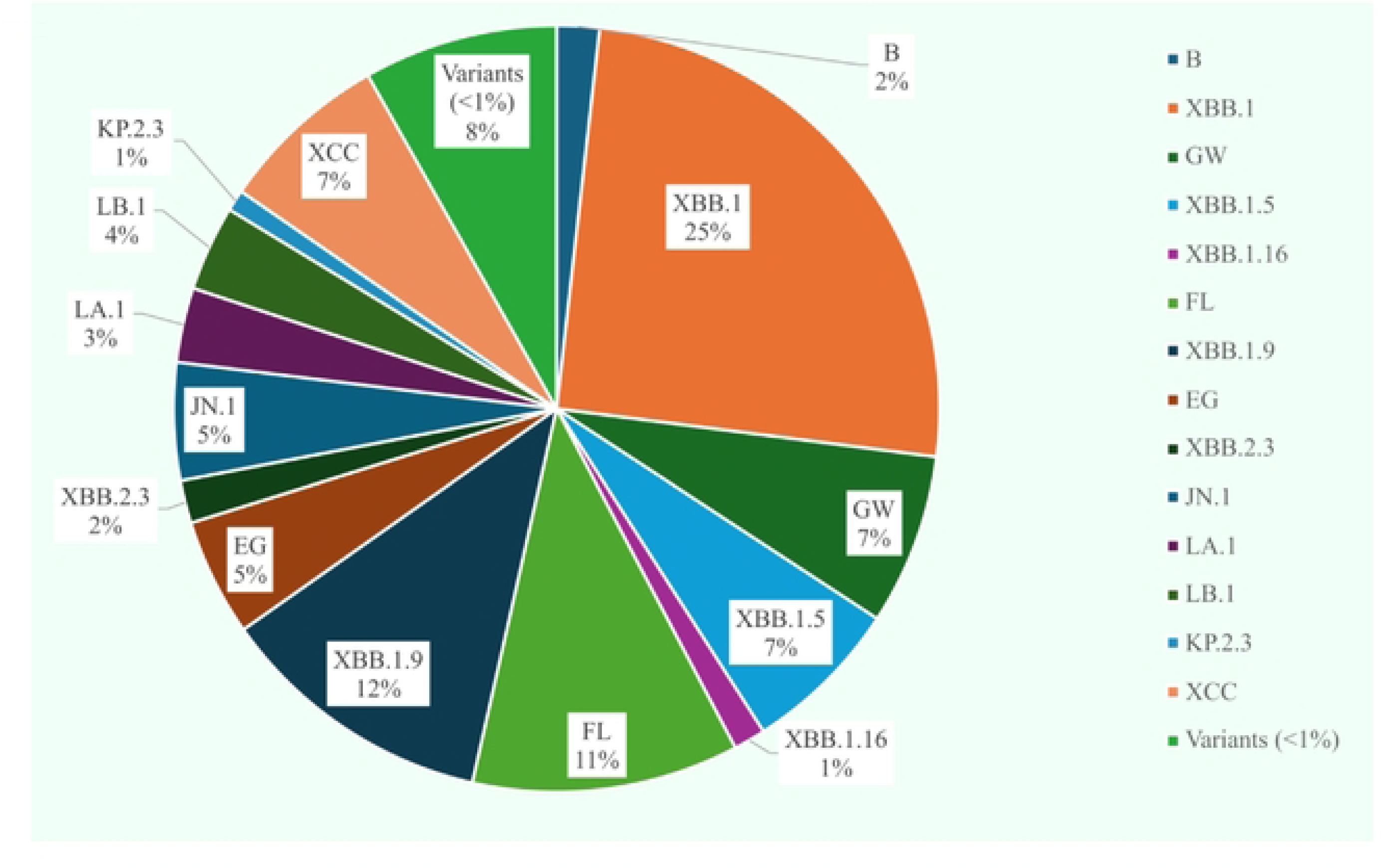
The pie chart illustrates the proportion of different SARS-CoV-2 variants (N=223). The major variant (≥5%) are XBB.1 (25%) shown in red color, XBB.1.9 (12%) grey, FL (11%) green, GW (7%) light green, XBB.1.5 (7%) light blue and EG (5%) dark yellow. The category “Other variants” less than 1% (11%) includes lineages detected at lower frequencies. The frequency of the respective number of lineages are given within parentheses

A heat map (**Fig 4)** depicts the month-wise frequency of SARS-CoV-2 variants. In the first quarter of 2023 the most prevalent variants included, XBB.1 (37%), XBB.1.9 (19%), FL (16%). Other lineages included BQ, BN, XBB, XBB.1.5 were 11%, B, EG, GW were 5% respectively. While the recombinant variant was 10% and XBB.2.4 (1%). In the first quarter of 2023, XBB.1 was observed to be 37%, which declined to 22% in the third quarter of 2023, FL lineages decreased to 17% from 36% in the second quarter of 2023, while EG lineage increased to 18% during the same period. The XBB.1.5 and XBB.1.9 variants both decreased to 9% in second quarter of 2023, XBB.2.3 also decreased to 8% during the same period. The XBB.1.16 lineage was also observed to be 9% in the second quarter of 2023 and increased to 11% in the third quarter of 2023, GW lineage was pre-dominant (100%) in the third quarter of 2023, however declined to 20% in the first quarter of 2024. Moreover, LA.1 variant was pre-dominant which increased from 40% in the first quarter of 2024 to 100% prevalence in the second quarter of 2024, the recombinant variant also showed 100% prevalence in the third quarter of 2024. LB.1 also showed a gradual increase, rising from 25% to 33% in the third quarter of 2024. Of note, the JN variant frequency decreased from 80% in the first quarter to 13% in the third quarter of 2024, respectively.

**Fig 4:**
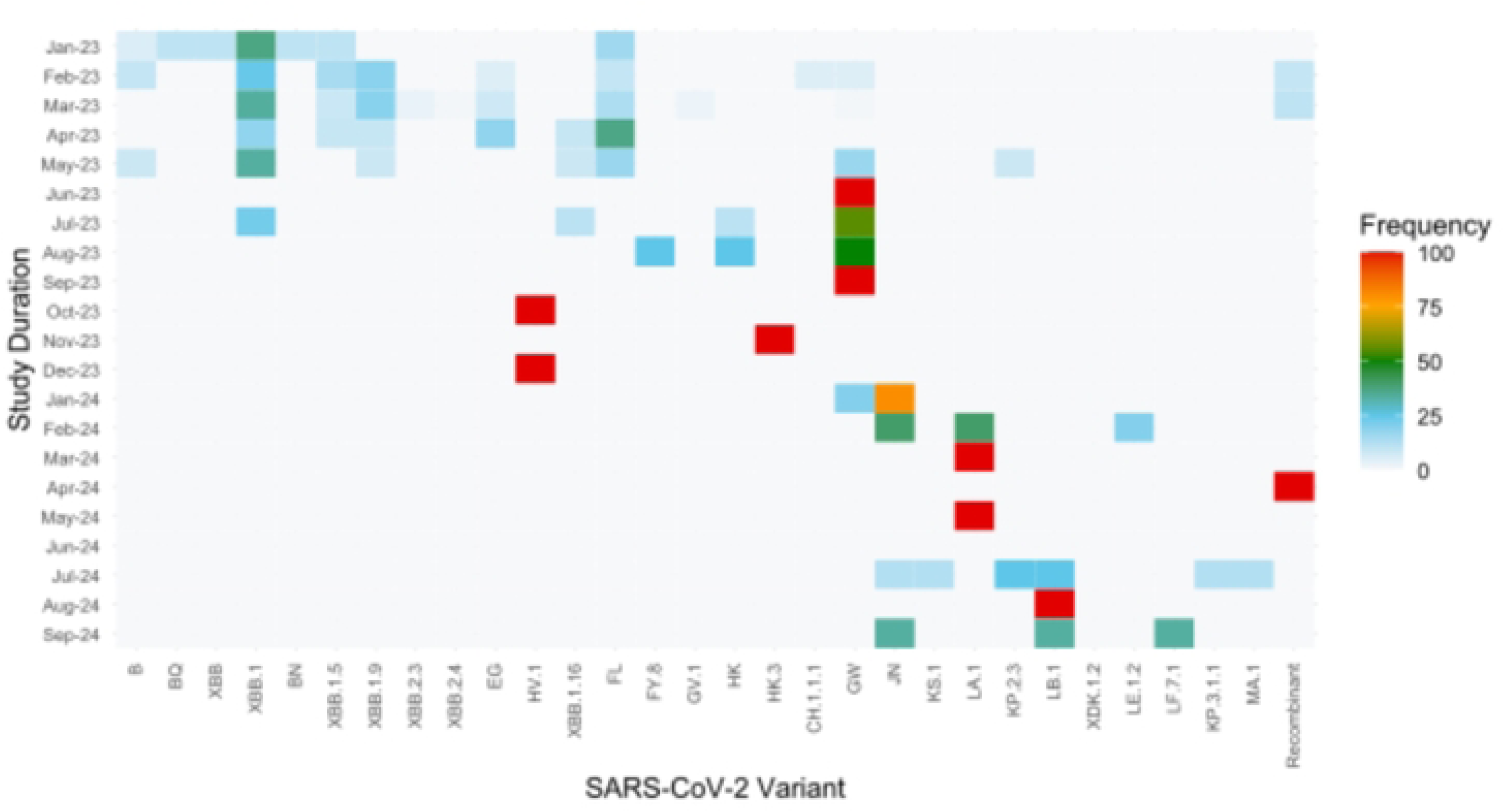
Heatmap illustrating the distribution of SARS-CoV-2 lineages and sublineages in Pakistan from January 2023 to September 2024. The frequency of each lineage is represented by a color gradient, where white color indicates a frequency of <5%, green represents frequencies between 5% and 50% (XBB.1, XBB.1.9, XBB.1.5, GW and FL) and red denotes frequencies >50% (HV.1, HK.3, GW, LA.1, LB and recombinant). The intensity of the colors corresponds to the relative proportion of each variant over time.

### Phylogenetic Analysis of SARS-CoV-2 and their evolutionary rates

A time-resolved maximum likelihood phylogenetic tree was constructed to examine the evolutionary dynamics of SARS-CoV-2 variants in Pakistan from September 2021 to August 2024. The sequences clustered into multiple clades including 21L, 22F, 23D, 23I, 24A, and 24H, reflecting lineage diversification over time. Clade 22F was dominant in mid-2022 and was gradually replaced by clade 23D and 23I variants in early to mid-2023. A sharp emergence of clade 24 variants, specifically 24A, 24H, and 24F clades were observed between late 2023 and August 2024. A total of 47 lineages, including XBB.1.9.1, JN.1, and KP.3.1.1, were identified, with increasing diversity toward the latter half of the surveillance period. This transition suggests ongoing viral evolution and local circulation of newer Omicron-derived sub variants (**Fig 5A-B**).

**Fig 5:**
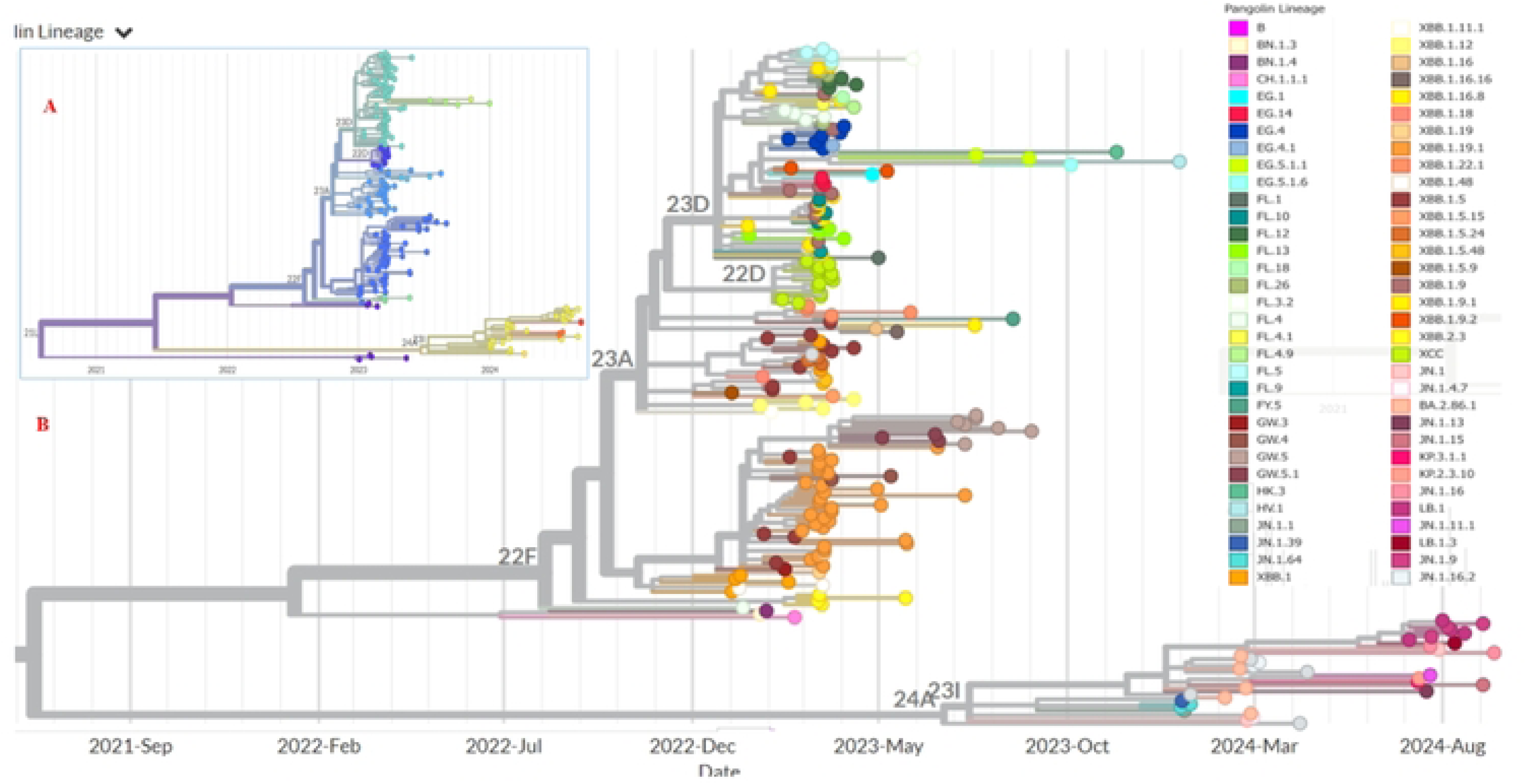
Phylogenetic tree of SARS-CoV-2 clades and Pangolin lineages in Pakistan (January 2023-September 2024). The phylogenetic tree represents the evolutionary relationships of SARS-CoV-2 whole genome sequences (n=215) from different regions of Pakistan collected between January 2023 and September 2024. **A.** The phylogenetic tree shows 14 distinct clades were present from January 2023 to September 2024. **B.** The phylogenetic analysis using augur pipeline showed 11 SARS-CoV-2 clades representation labelled on branches and nodes are highlighted by distinct colors, showing lineages emerging in Pakistani population with time. Variants of interest (VOC): XBB, JN.1, GW.5, LB.1, and emerging variants, are highlighted.

To quantify the evolutionary rate of SARS-CoV-2, a root-to-tip regression analysis was performed. A strong temporal signal was observed with a substitution rate estimate of 8.01 × 10⁻⁴ substitutions per site per year. The divergence pattern showed progressive accumulation of mutations over time, aligning with the introduction and dominance of clades 23D, 24A, and 24H The linear relationship between divergence and sampling date supports the molecular clock assumption and validates the reliability of the phylogenetic inferences (**Fig 6A-B**)

**Fig 6:**
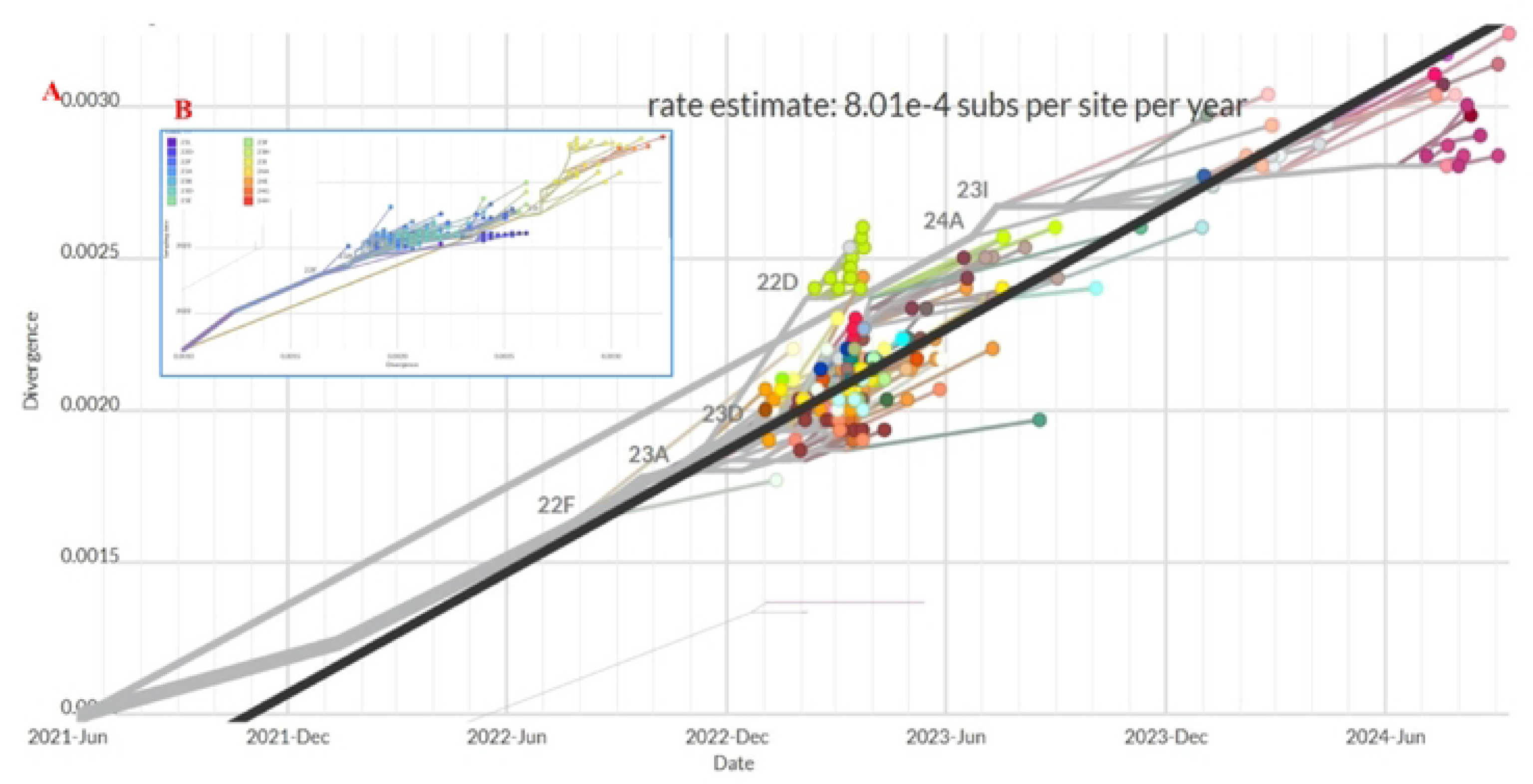
Genomic Divergence in SARS-CoV-2 Strains from January 2023 till September 2024 in Pakistan. **A.** Strong temporal signal with a substitution rate estimate of 8.01 × 10⁻⁴ substitutions per site per year. **B.** The divergence pattern showed progressive accumulation of mutations over time, aligning with the introduction and dominance of clades 23D, 24A, and 24H

### Comparison with global circulation

For global comparative analysis, SARS-CoV-2 genomic surveillance data were retrieved from the GISAID EpiCoV database for six additional countries with an aim to include data across geographical settings where greater surveillance information was present. Hence, we studied SARS-CoV-2 genomes reported from United States, United Kingdom, France, Germany, Italy, and India between 2023 and 2024. For each of these countries we depict frequently reported variants in 6 monthly intervals during the study period **(Fig 7)**. The total number of sequences retrieved per country were as follows: United States (n = 286,135), United Kingdom (n = 54,574), France (n = 32,137), Germany (n = 22,451), Italy (n = 12,509), and India (n = 14,734). For each country, the top 20 most frequently reported lineages during the study period were selected. It was evident that between January to June 2023, XBB.1.5 emerged as the dominant global variant, specifically in the United States (84%) by April 2023. Concurrently, XBB.1.16 and XBB.1.9 showed significant circulation in India and parts of Europe. By mid to late 2023 (July–December), the emergence of the JN.1 variant led to a rapid global expansion, representing over 60% of sequences in the UK by December 2023 and contributing to approximately 27% of global cases by late November 2023. By early 2024, JN.1 and its subvariants such as JN.1.1, KP.2, KP.3.1.1, and LB.1 became the predominant circulating variants globally, with several countries reporting over 70% prevalence of these variants. Of note, whilst KP.3.1.1 had become the predominant variant in USA, UK, Italy, France and Germany by September 2024, this was not the case in India where JN.1.1. remained predominant at this time. Referring back to the trends in Pakistan (**Fig 1)**, JN.1 and KP.2.3 were prominent by September 2024 (this study).

**Fig 7:**
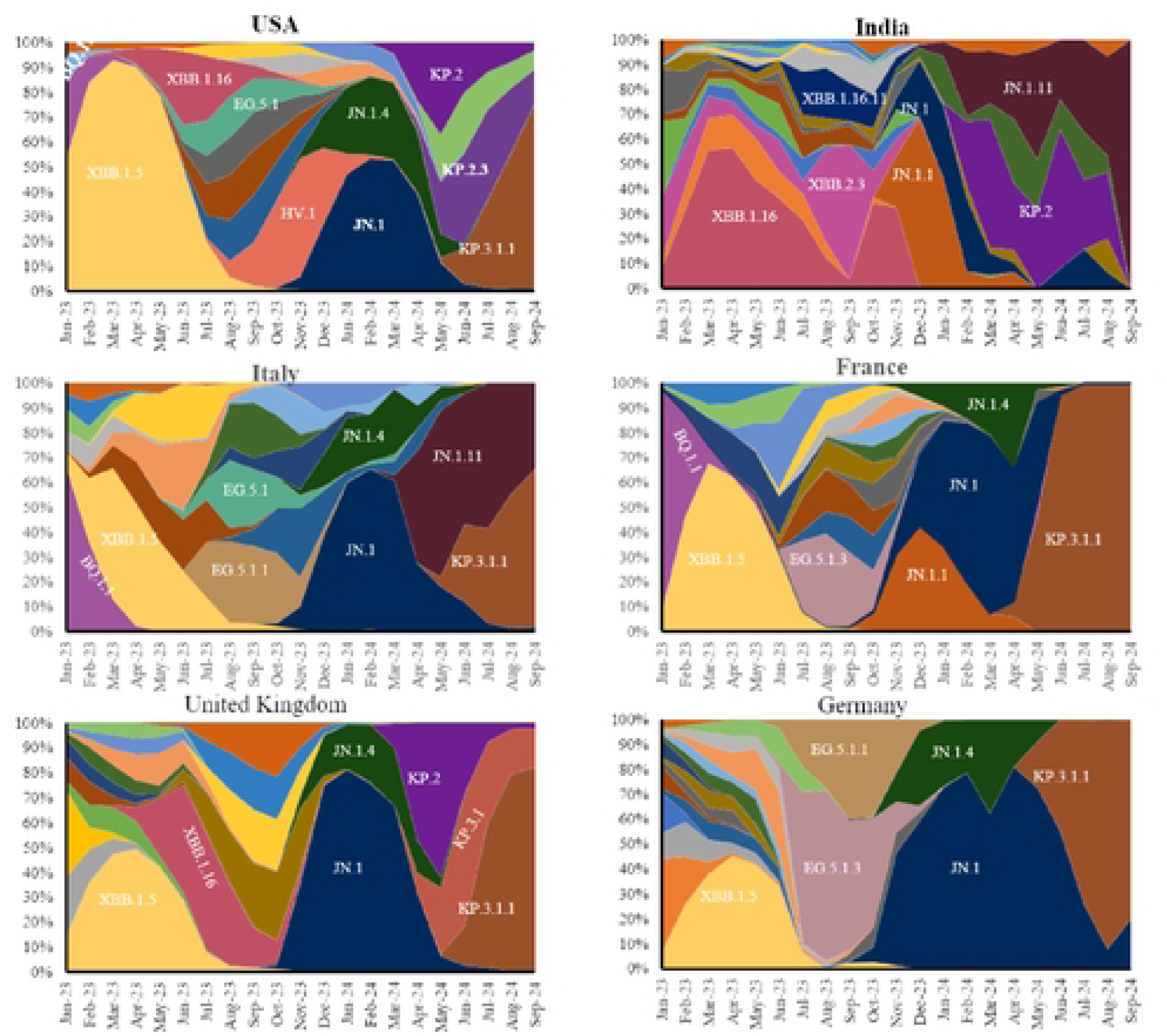
Area plots showing the temporal dynamics of the major SARS-CoV-2 variants and subvariants in the USA, UK, Germany, France, Italy, and India from January 2023 to September 2024. The plots represent the relative proportion of each variant detected per month based on data from GISAID. Major globally circulating lineages during this period included XBB.1.5, JN.1, XBB.1.16, and KP variant

## Discussion

The post-pandemic evolution of SARS-CoV-2 has been characterized by the emergence and diversification of multiple variants, driven by a combination of viral replication dynamics, host immune pressures, and vaccination-induced selection.

We studied the diversity of circulating SARS-CoV-2 PANGOLIN lineages and GISAID clades in Pakistan compared with global data for the period January 2023 and September 2024. Our data reveals a dynamic shift in variant predominance in Pakistan from early 2023 beginning, with XBB and its sub-variants and transitioning into FL, GW, and eventually JN.1 and HV.1 variants. This shift mirrors global trends in viral evolution under immune pressure and widespread vaccination (5, 22). Importantly, comparison with global data revealed differences in the emergence of variants between global data compared with Pakistan strains where JN.1 variants remaining dominant until September 2024.

In early 2023, the XBB variants such as XBB.1, XBB.1.9 and XBB.1.5 were pre-dominant in Pakistan. This trend was also observed worldwide and also in India, where similar subvariants showed increased transmissibility and significant immune evasion capabilities(23, 24). Also, with genomic surveillance reports from neighboring countries, particularly India and Southeast Asia (25). These variants, derived from recombination of BA.2 sublineages, were associated with increased transmissibility and moderate immune escape capabilities(26). The replacement of XBB by FL and GW sublineages by mid-2023 is also consistent with global observations, particularly from North America and Europe, where FL.1.5.1 and GW.5 rapidly gained ground (27, 28).Over time, the viral landscape has been shown to be diversified through recombinant forms like XCC and XDK.1.2, indicative of active viral evolution and adaptation under immune pressure (29).

The emergence of JN.1 and HV.1 as dominant variants in late 2023 and 2024 is particularly notable. JN.1, a descendant of BA.2.86, demonstrated significant growth advantage and immune evasion properties, becoming globally predominant by early 2024(24). Similarly, HV.1 emerged as a fast-spreading lineage with mutations in spike protein regions associated with decreased neutralization by vaccine-induced antibodies(30). Worldwide, there was a sharp increase in JN.1 variant in late 2023. Studies from UK Health Security Agency (UKHSA) and the Center of Disease Control (CDC) indicate JN.1 gained significant growth advantage. An editorial summarizing GISAID data reported approximately 27% global share of JN.1 sequences by late November 2023. Moreover, this variant showed a significant increase in France (20%) and the U.S (14%)(31). The SARS-CoV-2 variant, KP.2 is a descendant of JN.1 carrying key spike mutations, emerged by mid-2024 and was associated with rising COVID detection in parts of Europe. JN.1 sub variants, such as KP.3.1.1 and LB.1, emerged in parallel and spread globally by mid-2024(32–35).

Most data was available for COVID-19 samples reported between January to April 2023, and for January, February, July and August in 2024. Limited testing during the latter period could be attributed to reduced disease severity of COVID-19 from Omicron variants and COVID-19 vaccinations leading to reduced symptomatic infections (36). Our data show that majority of COVID-19 cases were aged between 19 to 50 years and greater than 50 years of age. Deaths have previously been associated with older age groups (37). Moreover, previous findings have shown that the gender-based differences in SARS-CoV-2 sensitivity and outcomes, mainly due to biological and behavioral factors(38). Our analysis of SARS-CoV-2 genomes at our facility in Karachi revealed the presence of 66 distinct variants and sub-variants, classified into 14 major clades. The findings reveal dynamic shifts in variant prevalence and predominance of SARS-CoV-2 variants such as XBB.1, XBB.1.9, FL and GW variants across all age groups aligns with global observations of variant emergence and spread. The World Health Organization (WHO) has emphasized the importance of monitoring such variants due to their potential impact on transmissibility and public health(5) Gender-wise it was observed that females exhibited higher prevalence of XBB.1.5, LB and FL SARS-CoV-2 variants, however in the males increasing trends of XBB.1, XBB.1.9, JN and GW variants were observed. These trends may be due to increased social and occupational exposures in this demographic.

Most samples were from Karachi, Sindh, this may be due to the population of Karachi with approximately 20 million inhabitants. Moreover, higher number of samples from a particular city may be due to the extensive network of diagnostic laboratories in Karachi. The highest case numbers and deaths have been previously shown to be recorded in Sindh and Punjab, the most populous provinces in Pakistan. Sindh province accounted for 37% of all COVID-19 cases in Pakistan (https://www.worldometers.info/coronavirus/country/pakistan). It was reported that 81% of COVID-19 cases across Sindh were from Karachi, with a population of 20 million (https://www.sindhhealth.gov.pk/daily_situation_report)(8, 39–41).

The highest diversity of clades was present in Sindh and Islamabad Capital Territory and the least in Punjab province. Initial introductions of SARS-CoV-2 GH (B.1.255, B.1) and S (A) clades were associated with overseas travelers. Additionally, GH (B.1.255, B.1, B.1.160, B.1.36), L (B, B.6, B.4), V (B.4) and S (A) clades were transmitted locally These findings highlight the virus’s continued evolutionary adaptability in response to host immune pressures and increasing population-level immunity(42). Our age-stratified analysis shows that SARS-CoV-2 variants such as XBB.1, XBB.1.16, XBB.1.5, EG, JN.1 and recombinant were more frequently observed in adults above 18 years, whilst XBB.1.9, FL and GW variants in less than 19 years of age. While our dataset does not include clinical severity or vaccination status, this distribution may reflect differences in exposure risk, occupational mobility, or waning immunity in older populations(43). Previous studies to understand COVID-19 immunity those related to understanding seroprevalence of SARS-CoV-2 and investigations on host immunity (44, 45). However, these are the first data to show that the strains circulating in Pakistan during the post-pandemic period are different from other countries and the comparison with the Global North which suffered comparatively higher rates of COVID-19 morbidity and mortality are of consequence.

Our findings emphasize the importance of global genomic monitoring and data-sharing platforms, such as GISAID, in tracking emerging SARS-CoV-2 variants. The XBB.1.9, XBB.1.5, JN and KP.2.3 variants have been previously reported to have mutations in the spike protein, that enhances their ability to escape neutralization by vaccine induced antibodies(46). Previous studies by European Centre for Disease Prevention and Control (ECDC) and United States Centers for Disease Control and Prevention (UCDCP) have also shown the emergence of the SARS-CoV-2 variants, which suggests the importance of continuous genomic surveillance to detect and respond to emerging variants (31, 33, 47).

## Limitations

COVID-19 reported from Pakistan were limited compared with those from many high-resource settings due to the limited testing and reporting capacity in the country. Further, genomic surveillance has been limited to public health laboratories and a few academic university settings such as the Aga Khan University (39, 41, 48). Therefore, a relatively small number of SARS-CoV-2 genomes were available for analysis. However, by studying both data from Pakistan available in GISAID and also from our facility we are able to study demographic trends associated with variant circulation.

## Conclusions

Our genomic surveillance study of SARS-CoV-2 from January 2023 to September 2024 reveals a dynamic landscape of viral evolution, marked by the emergence of multiple variants and sub-variants across diverse clades, emphasizing the importance of continued genomic surveillance. Given the ongoing evolution of SARS-CoV-2 and the limitations of current vaccine strategies, there is an urgent need for continuous genomic monitoring, rapid data sharing, and the development of next-generation vaccines targeting conserved viral regions.

## Data Availability

All data are available with this submission.

## Acknowledgements

We thank the staff at the Section of Molecular Pathology, Aga Khan University Hospital, Karachi, Pakistan for providing information and support for SARS-CoV-2 test samples.

## Declarations

### Ethics approval and consent to participate

This study was approved by the Ethical Review Committee, The Aga Khan University (AKU). No need for consent as the data from online resource GISAID is used for the analysis.

### Consent for publication

Not applicable

### Data availability

All genome sequences and associated metadata have been submitted to the GISAID EpiCoV™ database and are accessible at: https://www.gisaid.org. These are also included within the manuscript

## Supplementary Information

**S1 Fig.** SARS-CoV-2 genome submissions in GISAID from each region of Pakistan across time. The graph shows all SARS-CoV2 sequences (n=223) genomes submitted from each region of Pakistan between January 2023 and September 2024. Sindh (light blue), ICT (orange), Punjab (grey), KPK (yellow), Baluchistan (blue), GB (green). On x-axis date in month-year format is mentioned and y-axis shows number of SARS-CoV2 genomes submitted in GISAID

## References

1. Zhu N, Zhang D, Wang W, Li X, Yang B, Song J, et al. A Novel Coronavirus from Patients with Pneumonia in China, 2019. N Engl J Med. 2020;382(8):727–33.

2. Cucinotta D, Vanelli M. WHO Declares COVID-19 a Pandemic. Acta Biomed. 2020;91(1):157–60.

3. WHO. WHO Coronavirus (COVID-19) Dashboard. Accessed June 25, 2025.

4. Emmanuel F, Hassan A, Ahmad A, Reza TE. Pakistan’s COVID-19 Prevention and Control Response Using the World Health Organization’s Guidelines for Epidemic Response Interventions. Cureus. 2023;15(1):e34480.

5. WHO. Tracking SARS-CoV-2 Variants. 2024.

6. Cascella M RM, Aleem A, et al. Features, Evaluation, and Treatment of Coronavirus (COVID-19) StatPearls [Internet] Treasure Island (FL) 2023

7. Pakistan. Go. COVID-19 Data Portal (Ministry of National Health Services). (Accessed June 25, 2025).

8. Nasir A, Aamir UB, Kanji A, Bukhari AR, Ansar Z, Ghanchi NK, et al. Tracking SARS-CoV-2 variants through pandemic waves using RT-PCR testing in low-resource settings. PLOS Glob Public Health. 2023;3(6):e0001896.

9. Ghanchi NK, Nasir A, Masood KI, Abidi SH, Mahmood SF, Kanji A, et al. Higher entropy observed in SARS-CoV-2 genomes from the first COVID-19 wave in Pakistan. PLoS One. 2021;16(8):e0256451.

10. Bukhari AR, Ashraf J, Kanji A, Rahman YA, Trovao NS, Thielen PM, et al. Sequential viral introductions and spread of BA.1 across Pakistan provinces during the Omicron wave. BMC Genomics. 2023;24(1):432.

11. Khan AA, Abdullah M, Khan R, Kazmi T, Sultan F, Aamir S, et al. Pakistan’s national COVID-19 response: lessons from an emergent response to the pandemic. Front Public Health. 2024;12:1379867.

12. Pillay SG, J; Tegally, H; Eduan Wilkinson, etal Illumina Nextera DNA Flex library construction and sequencing for SARS-CoV-2:Adapting COVID-19 ARTIC protocol v1. 2020.

13. Network A. hCoV-2019/nCoV-2019 Version 3 Amplicon Set.”

14. Illumina. MiSeq System Denature and Dilute Libraries Guide

15. Huddleston J, Hadfield J, Sibley TR, Lee J, Fay K, Ilcisin M, et al. Augur: a bioinformatics toolkit for phylogenetic analyses of human pathogens. J Open Source Softw. 2021;6(57).

16. Katoh K, Misawa K, Kuma K, Miyata T. MAFFT: a novel method for rapid multiple sequence alignment based on fast Fourier transform. Nucleic Acids Res. 2002;30(14):3059–66.

17. Sagulenko P, Puller V, Neher RA. TreeTime: Maximum-likelihood phylodynamic analysis. Virus Evol. 2018;4(1):vex042.

18. Hadfield J, Megill C, Bell SM, Huddleston J, Potter B, Callender C, et al. Nextstrain: real-time tracking of pathogen evolution. Bioinformatics. 2018;34(23):4121–3.

19. Minh BQ, Schmidt HA, Chernomor O, Schrempf D, Woodhams MD, von Haeseler A, et al. IQ-TREE 2: New Models and Efficient Methods for Phylogenetic Inference in the Genomic Era. Mol Biol Evol. 2020;37(5):1530–4.

20. Organization WH. Statement on the fifteenth meeting of the IHR Emergency Committee regarding the COVID-19 pandemic. 2023.

21. Worldometer. COVID-19 Coronavirus Pandemic – Pakistan. 2024.

22. Tao K, Tzou PL, Nouhin J, Gupta RK, de Oliveira T, Kosakovsky Pond SL, et al. The biological and clinical significance of emerging SARS-CoV-2 variants. Nat Rev Genet. 2021;22(12):757–73.

23. Arora P, Cossmann A, Schulz SR, Ramos GM, Stankov MV, Jack HM, et al. Neutralisation sensitivity of the SARS-CoV-2 XBB.1 lineage. Lancet Infect Dis. 2023;23(2):147–8.

24. Tamura T, Ito J, Uriu K, Zahradnik J, Kida I, Anraku Y, et al. Virological characteristics of the SARS-CoV-2 XBB variant derived from recombination of two Omicron subvariants. Nat Commun. 2023;14(1):2800.

25. Chen Y, Zhao X, Zhou H, Zhu H, Jiang S, Wang P. Broadly neutralizing antibodies to SARS-CoV-2 and other human coronaviruses. Nat Rev Immunol. 2023;23(3):189–99.

26. ECDC. SARS-CoV-2 variants of concern. 2025.

27. CDC. COVID Data Tracker: Variant Proportions. 2025.

28. WHO. Weekly epidemiological update on COVID-19 - 2024. Geneva: World Health Organization. 2024.

29. Jackson B, Boni MF, Bull MJ, Colleran A, Colquhoun RM, Darby AC, et al. Generation and transmission of interlineage recombinants in the SARS-CoV-2 pandemic. Cell. 2021;184(20):5179–88 e8.

30. S. A. Omicron: A variant of concern not a cause of panic. Journal of Advanced Pharmaceutical Technology & Research. 2022;13(2):138–40.

31. Ma KC, Castro J, Lambrou AS, Rose EB, Cook PW, Batra D, et al. Genomic Surveillance for SARS-CoV-2 Variants: Circulation of Omicron XBB and JN.1 Lineages - United States, May 2023-September 2024. MMWR Morb Mortal Wkly Rep. 2024;73(42):938–45.

32. Planas D, Staropoli I, Planchais C, Yab E, Jeyarajah B, Rahou Y, et al. Escape of SARS-CoV-2 Variants KP.1.1, LB.1, and KP3.3 From Approved Monoclonal Antibodies. Pathog Immun. 2024;10(1):1–11.

33. Karyakarte RP, Das R, Rajmane MV, Dudhate S, Agarasen J, Pillai P, et al. Appearance and Prevalence of JN.1 SARS-CoV-2 Variant in India and Its Clinical Profile in the State of Maharashtra. Cureus. 2024;16(3):e56718.

34. Karyakarte RP, Das R, Potdar V, Kulkarni B, Joy M, Mishra M, et al. Tracking KP.2 SARS-CoV-2 Variant in India and the Clinical Profile of KP.2 Cases in Maharashtra, India. Cureus. 2024;16(8):e66057.

35. Roucaute D. KP.2 variant in France. 2024.

36. Nisar MI, Ansari N, Malik AA, Shahid S, Lalani KRA, Chandna MA, et al. Assessing the effectiveness of COVID-19 vaccines in Pakistan: A test-negative case-control study. J Infect. 2023;86(5):e144–e7.

37. Nasir N, Habib K, Iffat K, Khan N, Muhammad ZA, Mahmood SF. Clinical characteristics and outcomes of COVID-19: Experience at a major tertiary care center in Pakistan. J Infect Dev Ctries. 2021;15(4):480–9.

38. Hoang VT, Colson P, Levasseur A, Delerce J, Lagier JC, Parola P, et al. Clinical outcomes in patients infected with different SARS-CoV-2 variants at one hospital during three phases of the COVID-19 epidemic in Marseille, France. Infect Genet Evol. 2021;95:105092.

39. Umair M, Ikram A, Salman M, Haider SA, Badar N, Rehman Z, et al. Genomic surveillance reveals the detection of SARS-CoV-2 delta, beta, and gamma VOCs during the third wave in Pakistan. J Med Virol. 2022;94(3):1115–29.

40. Tamim S, Trovao NS, Thielen P, Mehoke T, Merritt B, Ikram A, et al. Genetic and evolutionary analysis of SARS-CoV-2 circulating in the region surrounding Islamabad, Pakistan. Infect Genet Evol. 2021;94:105003.

41. Nasir A, Bukhari AR, Trovao NS, Thielen PM, Kanji A, Mahmood SF, et al. Evolutionary history and introduction of SARS-CoV-2 Alpha VOC/B.1.1.7 in Pakistan through international travelers. Virus Evol. 2022;8(1):veac020.

42. Nyberg Tea. A standardised protocol for relative SARS-CoV-2 variant severity assessment, applied to Omicron BA.1 and Delta in six European countries Euroroundup, 28(36). 2023.

43. Viana R, Moyo S, Amoako DG, Tegally H, Scheepers C, Althaus CL, et al. Rapid epidemic expansion of the SARS-CoV-2 Omicron variant in southern Africa. Nature. 2022;603(7902):679-86.

44. Masood KI, Qaiser S, Abidi SH, Khan E, Hussain A, Ghous Z, et al. Dynamics of IgG Antibody Responses to SARS-CoV-2 Reveals Insight into Immunity During the Early Pandemic Period in Pakistan. SSRN; 2022.

45. Iqbal J, Hasan Z, Habib MA, Malik AA, Muhammad S, Begum K, et al. Evidence of rapid rise in population immunity from SARS-CoV-2 subclinical infections through pre-vaccination serial serosurveys in Pakistan. J Glob Health. 2025;15:04078.

46. Jian F, Feng L, Yang S, Yu Y, Wang L, Song W, et al. Convergent evolution of SARS-CoV-2 XBB lineages on receptor-binding domain 455-456 synergistically enhances antibody evasion and ACE2 binding. PLoS Pathog. 2023;19(12):e1011868.

47. Control ECfDPa. SARS-CoV-2 variants of concern 2025.

48. Ghanchi NK, Nasir A, Masood KI, Abidi SH, Mahmood SF, Kanji A, et al. Higher entropy observed in SAR-CoV-2 genomes from the first COVID-19 wave in Pakistan PLoS ONE. 2021;16(8)(e0256451):10.1371/journal.pone.0256451.

